# Food Insecurity During the First Year of COVID-19: An Analysis of Employment and Sociodemographic Factors Among a Longitudinal Cohort (CHASING COVID)

**DOI:** 10.1101/2022.09.20.22280094

**Authors:** Yvette Ng, Mindy Chang, McKaylee Robertson, Christian Grov, Andrew Maroko, Rebecca Zimba, Drew Westmoreland, Madhura Rane, Chloe Mirzayi, Angela M. Parcesepe, Sarah Kulkarni, William Salgado-You, Nevin Cohen, Denis Nash

## Abstract

**Objectives:** While much has been reported about the impact of COVID-19 on U.S. food insecurity, longitudinal data and the variability experienced by people working in different industries are limited. This study aims to further characterize individuals experiencing food insecurity during the pandemic in terms of employment and sociodemographic characteristics and degree of food insecurity.

**Methods:** The study sample consisted of people enrolled in a U.S. prospective cohort study (CHASING COVID) who completed all food insecurity questionnaires from Visit 1 (April-July 2020) through Visit 7 (May-June 2021). Descriptive statistics and logistic regression models were used to determine employment and sociodemographic correlates of food insecurity (using a screening question from the USDA HFSS). Patterns of food insecurity and utilization of food benefit programs were also examined.

**Results:** Thirty-one percent (1251/4019) of the sample were food insecure. Black and Hispanic respondents, households with children, and those with lower income and education levels had a higher odds of food insecurity. People employed in construction, leisure/hospitality and trade/transportation industries had the highest burden of both food insecurity and income loss. Among those reporting food insecurity, 40% were persistently food insecure (≥4 consecutive visits), and 46% did not utilize any food benefit programs.

**Conclusions:** The pandemic resulted in widespread food insecurity in our cohort, much of which was persistent. In addition to addressing sociodemographic disparities, future policies should focus on the needs of those working in vulnerable industries and ensure those experiencing food insecurity can easily participate in food benefit programs for which they are eligible.

## Introduction

Much research has been published which indicates that food insecurity rose in 2020 due to the widespread economic disruption during the COVID-19 pandemic.^1-7^ In contrast, the reported percentage of U.S. households experiencing food insecurity in 2020, as defined by the Household Food Security Survey (HFSS) data from the United States Department of Agriculture (USDA) was 10.5%, which is unchanged from 2019.^8^ Examining the USDA data by race/ethnicity, the percentage food insecure decreased for White non-Hispanic households, while it increased for Black non-Hispanic and Hispanic households from 2019 to 2020. This had the effect of maintaining the overall food insecurity rate at 10.5%, while widening the difference between Black non-Hispanic and White households, where Black non-Hispanic household experienced food insecurity at approximately three times the rate of White households in 2020. This widening disparity has been observed in other studies.^1, 9-13^

The federal government passed legislation including the Families First Coronavirus Response Act (FFCRA) and the Coronavirus Aid, Relief and Economic Security Act (CARES) in March 2020 to help provide economic relief by increasing unemployment benefits, providing direct payments (“stimulus checks”) to eligible individuals, and increasing support for child nutrition programs and the Supplemental Nutrition Assistance Program (SNAP).^14,15^ Given the widening racial/ethnic disparities, not everyone may have benefited equally from this additional legislation. It is critical to further understand and characterize the experience of those facing food insecurity during the pandemic to address their needs more effectively and to implement policy that can better serve individuals who are most in need in future crises.

Public health measures enacted to prevent spread of COVID-19 had a huge impact on businesses and employment, resulting in job losses or decreased income.^16-20^ The leisure/hospitality sector accounted for 39% of all job losses from February to April 2020 due to business closures required by COVID mitigation policies. Although this industry has recovered to some extent, there were 1.7 million fewer jobs in July 2021 compared to January 2020.^18^ U.S. households continue to experience economic hardship and income loss due to unemployment across many industries. Data are limited on the longitudinal view of how and among whom the rates of food insecurity changed over the course of the first year of the COVID pandemic, and how people were faring at the end of the first pandemic year. Given the limited longitudinal data, we lack a clear understanding of whether people who experienced food insecurity during the COVID pandemic experienced temporary or more persistent food insecurity throughout the year and to what extent those who are food insecure utilized food support programs. The objective of this study was to characterize the level of food insecurity and participants who experienced food insecurity during the COVID-19 pandemic in the U.S. This study adds to the literature by examining occupational data, income loss, and patterns of food insecurity in a large, diverse U.S. cohort over a time span of approximately one year.

## Methods

This study utilized data from the CHASING COVID Cohort study, a prospective U.S. national study of 6,740 adults (18 years of age or older). Online recruitment began in March 2020, and once enrolled, study participants completed online questionnaires (a “visit”) every one to three months. Additional details on eligibility and enrollment have been previously described.^21^ The analytic population was restricted to the N=4,019 participants who had completed all food security questions though the first year of the pandemic, from Visit 1 (V1, April-July 2020) through Visit 7 (V7, May-June 2021).

### Study Variables

#### Food insecurity

The following question from the USDA’s HFSS was asked at each visit beginning with Visit 1 using a 30-day reference period: *The food that we bought just didn’t last, and we didn’t have money to get more* (response options: ‘often true,’ ‘sometimes true,’ or ‘never true’). An affirmative answer (‘often true’ or ‘sometimes true’) is an appropriate screen for food insecurity as detailed in prior literature.^22^ Participants were dichotomized into two categories: food insecure (response of ‘often true’ or ‘sometimes true’ at any visit, V1-V7), or food secure (response of ‘never true’ at every visit, V1-V7).

#### Food insecurity patterns

To understand patterns of food insecurity, the following three patterns were examined: 1) the number of assessments (out of 7 total) in which each participant reported being food insecure, 2) newly food insecure (participants reporting food insecurity for the first time during the study), and 3) persistent or temporary food insecurity. A participant was considered temporarily food insecure if they reported food insecurity in 3 or fewer consecutive visits and considered persistently food insecure if they reported food insecurity in 4 or more consecutive visits. These cutoffs were based on the median number of visits participants in the food insecure group experienced food insecurity.

#### Food support programs

To understand to what extent participants were using various food support programs, the following question was included at Visit 6 : *Since the pandemic began, have you used any of the following*? (response options: ‘food pantry,’ ‘soup kitchen,’ ‘SNAP,’ ‘Pandemic EBT,’ ‘emergency food support,’ ‘Other food support not listed above,’ ‘none’). Multiple responses were permitted. SNAP refers to the federal Supplemental Nutrition Assistance Program, and Pandemic EBT (P-EBT) refers to the federal program implemented in response to the pandemic which provided funds to children who would normally receive free or reduced meals in schools.

#### Income loss

Participants were asked at each visit beginning with Visit 1: *In the past month, have you experienced a significant personal loss of income as a result of the new coronavirus?* (response options: ‘Yes,’ ‘No,’ or ‘NA’). Respondents were considered to have ever experienced income loss if they responded yes at any visit to this question.

#### Occupation / Industry

Occupational data were collected at Visit 6. Respondents were asked to select one occupation from a list of 30+ choices that best described their job. These choices were largely based on the North American Industry Classification System (NAICS) used in the Census. These occupations were then categorized into one of the eleven NAICS Supersectors, hereafter referred to as the industry variable.^23^

#### Sociodemographic Variables

Participants reported other sociodemographic variables (age and gender) at Visit 1.

### Statistical analysis

Descriptive statistics were generated to examine the relationship between food security status groups (main outcome of interest) and sociodemographic variables (age, gender, race/ethnicity, education, employment, and income), presence of children in the household, region of residence (grouped into 4 regions as defined by the U.S. Census Bureau,^24^ and a 5^th^ region for U.S. territories), if area of residence is rural, urban or suburban, and reported income loss. Differences between the groups across variables of interest were assessed using chi-squared tests and post hoc analyses for significant chi-square values were carried out by calculating the significant adjusted residuals. To identify correlates of food insecurity, we ran a logistic regression model with the following independent variables: sociodemographics, presence of children in household, region of residence, rural/urban residence, and income loss. Variables found to be significant in these individual models were included in the final adjusted model. Differences in sociodemographic variables between the study cohort and the full CHASING COVID cohort were assessed using chi-squared tests and estimates from univariate and adjusted logistic regression models were compared qualitatively. The burden of food insecurity in the study cohort was further characterized by calculating per visit percent reporting food insecurity, percent newly reporting food insecurity and persistence of food insecurity. We used a scatter plot to display the percentage food insecure (Y) and percentage with any income loss (X) by industry type. Any p-value of < 0.05 was considered statistically significant. SAS version 9.4 (SAS Institute, Cary, NC) was used for all analyses.

## Results

### Cohort characteristics

Overall, 4,019 (60% of N=6740 enrolled into longitudinal follow-up) study participants completed the food insecurity question at each visit and were included in this analysis. Participants were enrolled from all 50 states, with 64.8% identifying as non-Hispanic White, 15.3% Hispanic, 9.4% non-Hispanic Black, 7.5% Asian/Pacific Islander, and 3.0% Other. 52.6% of participants identified as female, 44.7% male, and 2.8% non-binary. The majority of participants did not have children in the household (71.1%), were college graduates (65.9%), and were employed (62.4%). Descriptive statistics are presented in Table Differences between the full CHASING COVID cohort and the study cohort are noted in Appendix A.

### Characteristics of People Experiencing Food Insecurity

In total, 31.3% of the cohort responded at least once during the study that the food they bought just didn’t last and they didn’t have money to get more, and are thus categorized as food insecure. Food insecurity varied widely based on race and ethnicity, with 57% of Black respondents and 49% of Hispanic respondents being food insecure compared to 27% of Asian/Pacific Islanders and 23% of White respondents. Food insecurity was more common among 18-29 year-olds, those with less than a high school education, income less than $35,000/year, and those reporting being out of work or being homemakers. Prevalence of food insecurity varied by region, with 40% seen in the South and 23% seen in the Northeast. The majority of those who were food insecure also reported having experienced a significant income loss as a result of the pandemic (69.9%).

Table 2 presents results from the crude and adjusted logistic regression models examining the relationship of food insecurity and various sociodemographic characteristics. In the adjusted model, Black (aOR=2.61, 95% CI 1.99-3.42) and Hispanic (aOR=1.93, 95% CI 1.56-2.41) participants had approximately two-fold increased odds of being food insecure compared to White participants. Households with children had 2.3 times (95% CI 1.95, 2.84) the odds of being food insecure compared to those without children. Those residing in areas considered rural had 1.22 times the odds (95% CI 1.00, 1.50) of being food insecure compared to those residing in urban areas. Those reporting significant income loss had 3.8 times the odds (95% CI 3.21, 4.52) of being food insecure compared to those who did not report income loss. The odds of experiencing food insecurity decreased with increasing age and education level. The estimates from the study cohort are directionally similar to the estimates from the full CHASING COVID cohort (Appendix B).

**Table 1.** Baseline demographics of overall cohort and those who are food secure and food insecure

|  | Food insecure,<br>No. (%) | Food secure,<br>No. (%) | Total,<br>No. (%) | P value<br>(chi sq) |
| --- | --- | --- | --- | --- |
| Total | 1251 (31.13) | 2768 (68.87) | 4019 (100.00) |  |
| Age group |  |  |  |  |
| 18-29 | 316 (25.26) | 501 (18.10) | 817 (20.33) | <0.001 |
| 30-39 | 427 (34.13) | 737 (26.63) | 1164 (28.96) |  |
| 40-49 | 262 (20.94) | 484 (17.49) | 746 (18.56) |  |
| 50-59 | 140 (11.19) | 423 (15.28) | 563 (14.01) |  |
| 60+ | 106 (8.47) | 623 (22.51) | 729 (18.14) |  |
| Gender |  |  |  |  |
| Male | 538 (43.01) | 1257 (45.41) | 1795 (44.66) | 0.31 |
| Female | 680 (54.36) | 1433 (51.77) | 2113 (52.58) |  |
| Non-binary | 33 (2.64) | 78 (2.82) | 111 (2.76) |  |
| Race |  |  |  |  |
| Hispanic | 299 (23.90) | 314 (11.34) | 613 (15.25) | <0.001 |
| Black Non-Hispanic | 216 (17.27) | 161 (5.82) | 377 (9.38) |  |
| Asian/Pacific Islander Non-Hispanic | 80 (6.39) | 221 (7.98) | 301 (7.49) |  |
| White Non-Hispanic | 592 (47.32) | 2014 (72.76) | 2606 (64.84) |  |
| Other Non-Hispanic | 64 (5.12) | 58 (2.10) | 122 (3.04) |  |
| Children in household |  |  |  |  |
| No | 716 (57.23) | 2183 (77.42) | 2859 (71.14) | <0.001 |
| Yes | 535 (42.77) | 625 (22.58) | 1160 (28.86) |  |
| Education level |  |  |  |  |
| Less than HS | 46 (3.68) | 9 (0.33) | 55 (1.37) | <0.001 |
| HS | 200 (15.99) | 137 (4.95) | 337 (8.39) |  |
| Some college | 442 (35.33) | 535 (19.33) | 977 (24.31) |  |
| College grad | 563 (45.00) | 2087 (75.40) | 2650 (65.94) |  |
| Employment status |  |  |  |  |
| Employed | 730 (58.35) | 1776 (64.16) | 2506 (62.35) | <0.001 |
| Out of work | 238 (19.02) | 255 (9.21) | 493 (12.27) |  |
| Homemaker | 112 (8.95) | 109 (3.94) | 221 (5.50) |  |
| Student | 98 (7.83) | 212 (7.66) | 310 (7.71) |  |
| Retired | 73 (5.84) | 416 (15.03) | 489 (12.17) |  |
| Income |  |  |  |  |
| <\$35K | 585 (46.76) | 459 (16.58) | 1044 (25.98) | <0.001 |
| \$35-49,999 | 200 (15.99) | 256 (9.25) | 456 (11.35) | |
| \$50-69,999 | 164 (13.11) | 435 (15.72) | 599 (14.90) | |
| \$70-99,999 | 145 (11.59) | 565 (20.41) | 710 (17.67) | |
| \$100K | 157 (12.55) | 1053 (38.04) | 1210 (30.11) | |
| Region |  |  |  |  |
| Northeast | 276 (22.06) | 914 (33.02) | 1190 (29.61) | <0.001 |
| Midwest | 237 (18.94) | 487 (17.59) | 724 (18.01) |  |
| South | 447 (35.73) | 676 (24.42) | 1123 (27.94) |  |
| West | 291 (23.26) | 687 (24.82) | 978 (24.33) |  |
| US territories | 0 (0.00) | 4 (0.14) | 4 (0.10) |  |
| Designation |  |  |  |  |
| Rural | 432 (34.53) | 760 (27.46) | 1192 (29.66) | <0.001 |
| Suburban | 311 (24.86) | 737 (26.63) | 1048 (26.08) |  |
| Urban | 508 (40.61) | 1271 (45.92) | 1779 (44.26) |  |
| Income loss |  |  |  |  |
| No | 376 (30.06) | 1876 (67.77) | 2252 (56.03) | <0.001 |
| Yes | 875 (69.94) | 892 (32.23) | 1767 (43.97) |  |

**Table 2.** Correlation of demographic factors with food insecurity

|  | Univariable Analysis |  | Multivariable Analysis |  |
| --- | --- | --- | --- | --- |
|  | OR (95% CI) | P value | OR (95% CI) | P value |
| Age group |  |  |  |  |
| 18-29 (ref) | - | - | - | - |
| 30-39 | 0.92 (0.7, 1.11) | <0.001 | 1.03 (0.81, 1.32) | <0.001 |
| 40-49 | 0.86 (0.70, 1.06) | <0.001 | 0.90 (0.68, 1.18) | 0.03 |
| 50-59 | 0.53 (0.41, 0.67) | 0.01 | 0.60 (0.44, 0.82) | 0.04 |
| 60+ | 0.27 (0.21, 0.35) | <0.001 | 0.40 (0.28, 0.58) | <0.001 |
| Gender |  |  |  |  |
| Male (ref) | - | - | - | - |
| Female | 1.11 (0.97, 1.27) | 0.35 | - | - |
| Non-binary | 1.00 (0.65, 1.50) | 0.76 | - | - |
| Race |  |  |  |  |
| NH white (ref) | - | - | - | - |
| Hispanic | 3.24 (2.70, 3.89) | <0.001 | 1.93 (1.56, 2.41) | 0.20 |
| Black | 4.56 (3.65, 5.71) | <0.001 | 2.61 (1.99, 3.42) | <0.001 |
| Asian/PI | 1.23 (0.94, 1.62) | <0.001 | 1.12 (0.81, 1.54) | 0.002 |
| Other | 3.75 (2.60, 5.42) | 0.001 | 2.55 (1.63, 3.97) | 0.03 |
| Children in household |  |  |  |  |
| No (ref) | - | - | - | - |
| Yes | 2.56 (2.22, 2.96) | <0.001 | 2.35 (1.95, 2.84) | <0.001 |
| Education level |  |  |  |  |
| Less than HS (ref) | - | - | - | - |
| HS | 0.29 (0.14, 0.60) | 0.04 | 0.23 (0.10, 0.55) | 0.35 |
| Some college | 0.16 (0.08, 0.33) | 0.003 | 0.20 (0.09, 0.46) | 0.02 |
| College grad | 0.05 (0.03, 0.11) | <0.001 | 0.11 (0.05, 0.25) | <0.001 |
| Employment status |  |  |  |  |
| Employed (ref) | - | - | - | - |
| Out of work | 2.27 (1.87, 2.76) | <0.001 | 1.05 (0.82, 1.34) | 0.31 |
| Homemaker | 2.50 (1.89, 3.30) | <0.001 | 1.27 (0.88, 1.83) | 0.04 |
| Student | 1.13 (0.87, 1.45) | 0.43 | 0.55 (0.40, 0.77) | <0.001 |
| Retired | 0.43 (0.33, 0.56) | <0.001 | 0.99 (0.67, 1.46) | 0.75 |
| Income |  |  |  |  |
| <\$35K (ref) | - | - | - | - |
| \$35-49,999 | 0.61 (0.49, 0.77) | <0.001 | 0.67 (0.52, 0.87) | <0.001 |
| \$50-69,999 | 0.30 (0.24, 0.37) | 0.12 | 0.33 (0.26, 0.43) | 0.05 |
| \$70-99,999 | 0.20 (0.16, 0.25) | <0.001 | 0.27 (0.21, 0.35) | <0.001 |
| \$100K | 0.12 (0.10, 0.14) | <0.001 | 0.17 (0.13, 0.22) | <0.001 |
| Region |  |  |  |  |
| Northeast (ref) | - | - | - | - |
| Midwest | 1.61 (1.31, 1.98) | 0.24 | 1.04 (0.81, 1.33) | 0.98 |
| South | 2.19 (1.83, 2.62) | <0.001 | 1.21 (0.97, 1.52) | 0.03 |
| West | 1.40 (1.16, 1.70) | 0.31 | 0.93 (0.73, 1.17) | 0.12 |
| Designation |  |  |  |  |
| Urban | - | - | - | - |
| Rural | 1.42 (1.22, 1.66) | <0.001 | 1.22 (1.00, 1.50) | 0.10 |
| Suburban | 1.06 (0.89, 1.25) | 0.12 | 1.09 (0.89, 1.34) | 0.89 |
| Income loss |  |  |  |  |
| No (ref) | - | - | - | - |
| Yes | 4.89 (4.24, 5.66) | <0.001 | 3.81 (3.21, 4.52) | <0.001 |

### Patterns of Food Insecurity

Although approximately one-third of participants reported being food insecure at any time during the first year of the pandemic, the percentage food insecure at any given visit was 15-19%, with the highest percentage observed at Visit 1 and declining after Visit 3 (Figure 1). Participants who experienced food insecurity did so at a median of 4 visits (IQR: 1,6), with 23% of the food insecure group reporting being food insecure at all 7 visits. Among participants who experienced food insecurity, 40% were persistently food insecure and 60% were temporarily food insecure. At all visits, there were participants considered newly food insecure.

**Figure 1:**
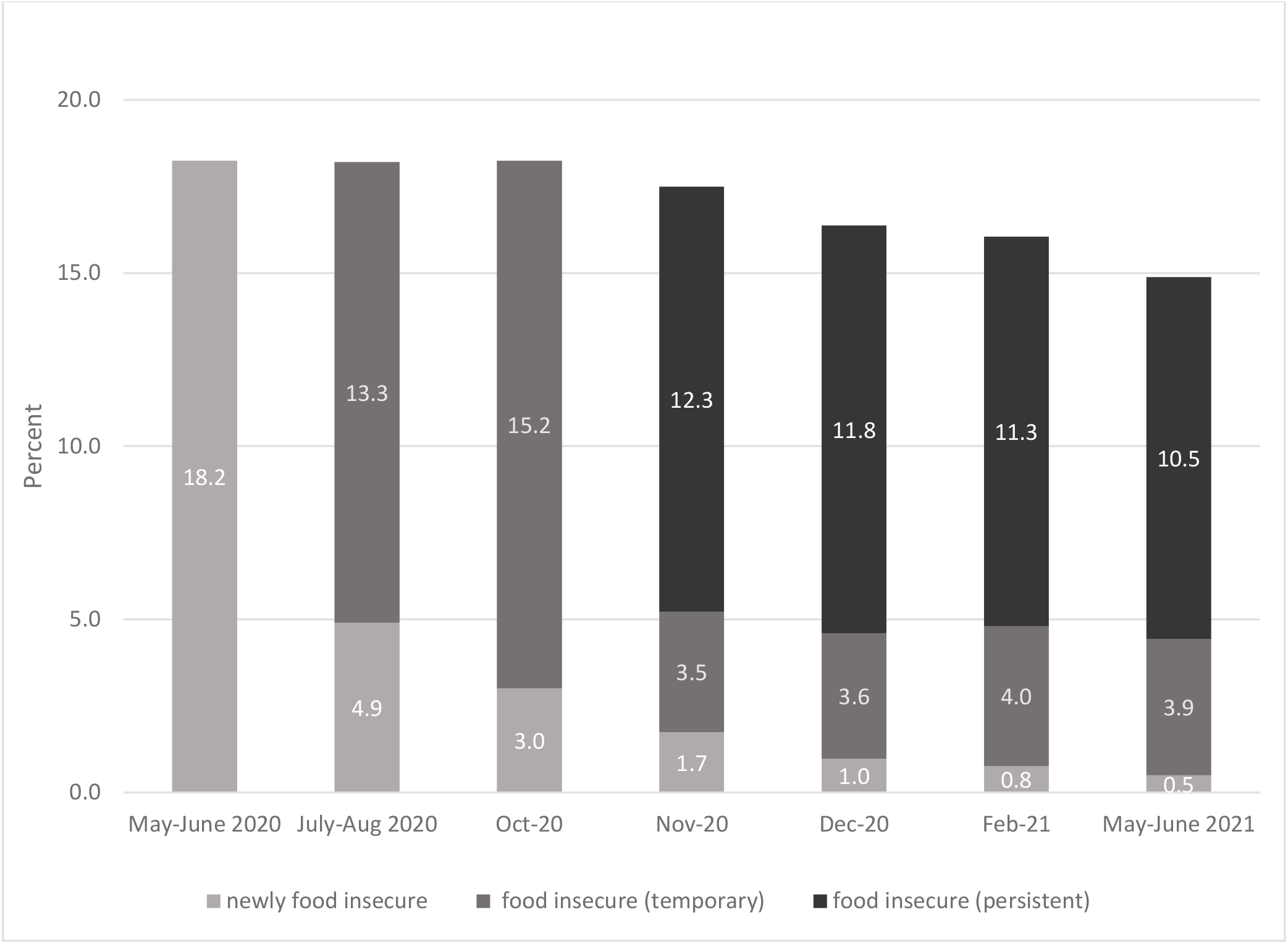
Percent food insecure by visit (and by level/category of food insecurity)

### Food Insecurity, Income Loss, and Industry/Occupation

As shown in Figure 2a, the percentage food insecure increased with percentage of any income loss due to the pandemic by industry type. Participants working in the construction, leisure/hospitality and trade/transportation industries had the highest percentages of persistent (29%, 19% and 19% respectively) and temporary food insecurity (25%, 25% and 24% respectively), whereas those in government had the lowest percentages (1% and 14% for persistent and temporary food insecurity respectively) (Figure 2b). Across all industries, 73-100% of those who reported persistent food insecurity and 48-84% of those who reported temporary food insecurity reported income loss at any visit.

**Figure 2:**
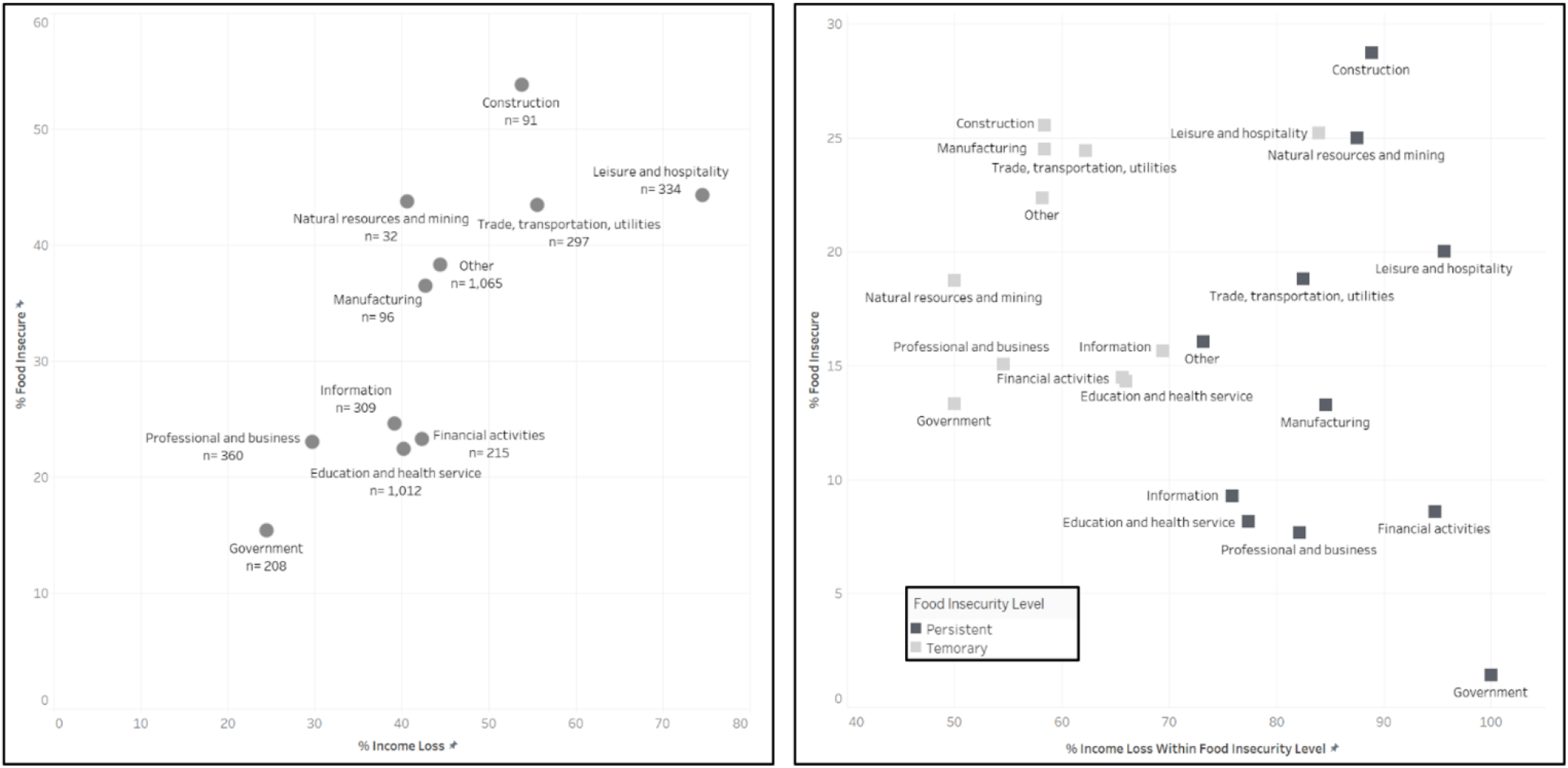
Food Insecurity, Income Loss, and Industry/Occupation Figure 2a: Percentage reporting income loss and percentage reporting food insecurity, by industry. Figure 2b: Percentage reporting income loss according to level of food insecurity, and percentage of each level of food insecurity, by industry. Note: Category of “Other” includes students, homemakers, disabled, religious, self-employed, other

### Use of Food Support Programs

To address their food needs, 75% of participants in the persistently food insecure group, 40% in the temporarily food insecure group, and 9% in the food secure group indicated using some form of food support program (Figure 3). The most common programs used were SNAP and food pantries, with those who were persistently food insecure reporting greater use of all food support programs.

**Figure 3:**
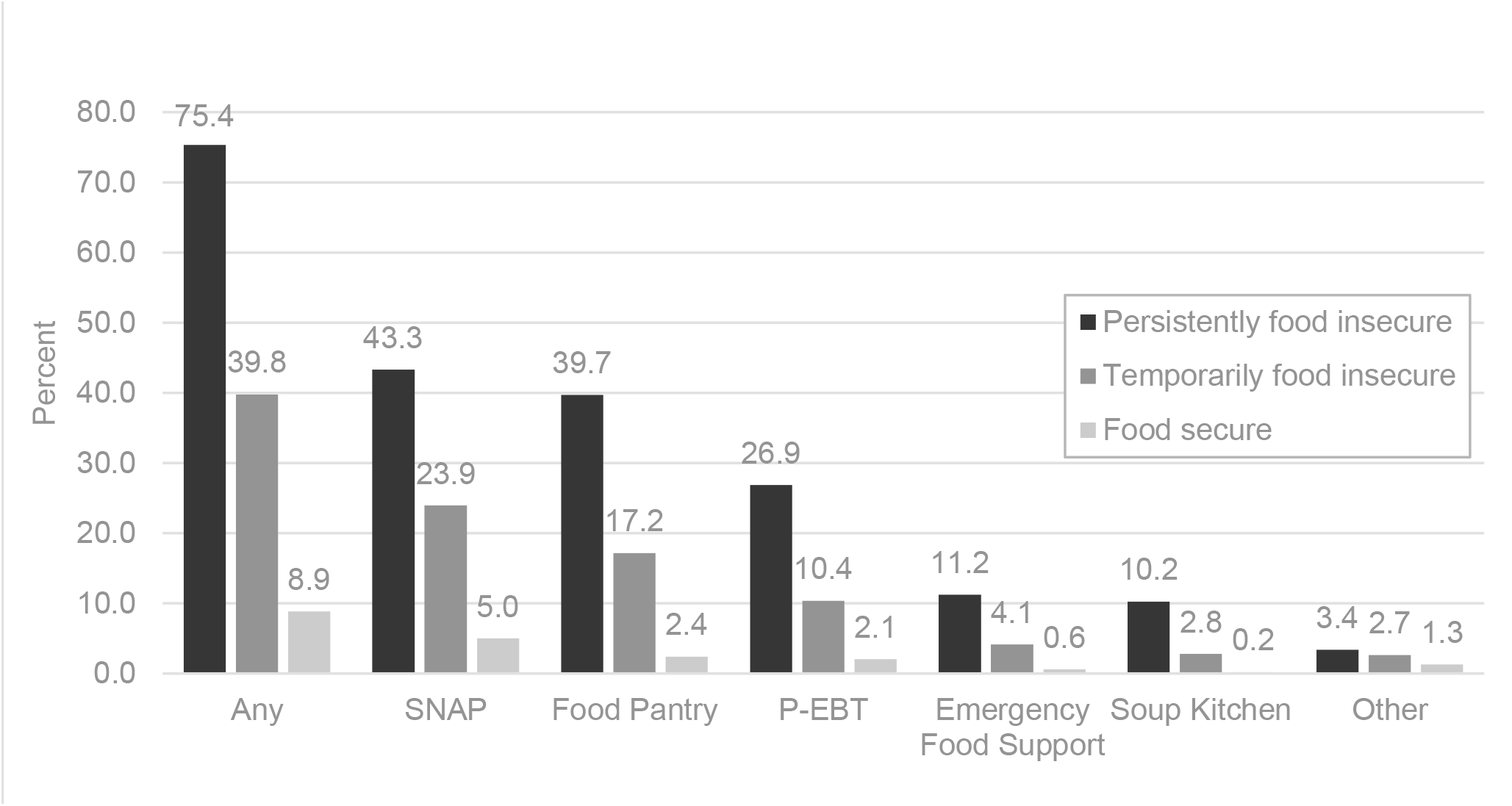
Utilization of Food Support Programs

## Discussion

This study adds new evidence to the literature on COVID’s impact on food insecurity by reporting not just overall prevalence of food insecurity but illuminating the extent to which people suffered from food insecurity. Almost one-third of our cohort was food insecure during the first year of the pandemic, consistent with existing reports indicating widespread food insecurity^,3,5,25-26^. What is notable from our longitudinal data is that 40% of those reporting food insecurity were persistently food insecure, indicating that for many people, there were prolonged difficulties in obtaining sufficient food and nutrition. In addition, this study shows that food insecurity was strongly related to income loss experienced as a result of the pandemic, and that there was wide variability seen across different industries.

Industries in which employees are not able to work remotely and not designated essential (construction, leisure/hospitality and trade/transportation) had the highest percentages of food insecurity. Unsurprisingly, sectors where participants had high percentages food insecurity also had high percentage with any income loss due to the pandemic, as seen in several other studies.^4,26,30-31^ Many occupations within these industries commonly have lower wages and minimal or no benefits such as sick leave and health care, further compounding the impact of the pandemic. A study based on the Census Household Pulse Survey found an inverse relationship between pre-pandemic income levels and financial hardship during the pandemic, which disproportionately affected Black and Hispanic Americans.^27^ The higher financial hardship seen among this population was associated with a greater risk of food insufficiency. Consistent with other research,^2,4,5,11,13,19,27-29^ our study showed that food insecurity during the pandemic disproportionately affected Black and Hispanic individuals/households, with approximately half of all Black and Hispanic participants reporting food insecurity (compared to 31% overall). Our data also indicated that Black and Hispanic respondents within the most affected industries tended to have lower incomes (as reported in Visit 1) than White respondents, which combined with lost income during the pandemic may have exacerbated the disparity.

To address food insecurity resulting from the pandemic, federal legislation like the FFCRA and the CARES Act expanded government support programs like SNAP (via expanded eligibility and increased benefits), implemented P-EBT, and provided funding for states to better support local food pantries. Among our cohort, a quarter of those who were persistently food insecure and more than half of those who were temporarily food insecure did not utilize any of these programs to alleviate their food insecurity, while only 43% of those persistently food insecure and 24% of those temporarily food insecure used SNAP. A study of Latino college students who became food insecure during the pandemic highlighted that a key barrier to food support programs was that they may not have met eligibility criteria or did not know how to access the resources.^32^ Studies conducted during the pandemic showing that less than half of those who lost employment income applied for unemployment benefits^27^ and that increases in food insecurity were not accompanied by similar increases in SNAP participation,^5^ suggesting that barriers to food assistance exist for those who are food insecure. Similarly, the level of assistance may not be adequate to prevent food insecurity. Forty percent of those in our study reporting participation in SNAP were persistently food insecure. Our data show participants experiencing food insecurity for the first time one year into the pandemic, indicating that current methods of addressing food insecurity are not sufficient and/or the impacts of the pandemic may not be felt by all immediately. The re-evaluation of the Thrifty Food Plan in August 2021, which increased monthly SNAP benefits an average of 21%, is a step in the right direction, but it is largely offset by the expiration of other benefit increases implemented during the pandemic.^33^ Further analyses using this dataset are planned to examine how availability and timing of various benefits (e.g., unemployment insurance, regional differences in policies) impacted food insecurity.

Our study has several limitations. Information collected to assess food insecurity did not consist of the fully validated USDA HFSS (6 or 18 question version), thus our data cannot be directly compared. A single question from the HFSS was used as a proxy to determine food insecurity as a binary variable, and USDA-defined levels of food insecurity (i.e., low, very low) could not be calculated. We did not collect information on food insecurity status prior to the start of the pandemic, therefore, we cannot assess if or by how much food insecurity changed as a result of the pandemic. We restricted our study to the 4,019 participants who completed the question at each visit as opposed to the 6,740 who completed the question at 1 or more visits, which could be a source of selection bias, but regression models using both groups produced similar findings. Finally, our approach to identifying correlates of food insecurity was based on a predictive model that includes all significant variables. The model identifies correlates and should not be used to inform causal inference. However, we believe the results of this descriptive study are useful for hypothesis generation. Strengths of our study include the composition of our study sample, from a large U.S. cohort (>4000 respondents) that is diverse among a variety of characteristics (race, age, geographic location). This study includes participants followed longitudinally for over one year over the course of the pandemic, with data captured at seven time points.

## Conclusions

Our study showed that a large proportion of people experiencing food insecurity during the first year of the pandemic did so on a persistent basis, and that certain populations, such as those working in vulnerable industries and those who experienced income loss, were much more likely to report food insecurity. In addition, a quarter of those who experienced persistent food insecurity did not seek assistance from either government or emergency food programs. The results illustrate the importance of targeted policies that help workers in industries most affected by pandemic-related income loss. It is also critical to publicize the availability of such assistance and make it easily accessible to those for whom it is intended. Policies and programs should focus on outreach to communities to ensure people are aware of the benefits for which they are eligible and entitled. Government agencies, working with community organizations as applicable, should minimize administrative and logistical burdens wherever possible to facilitate enrollment.

## Data Availability

All data produced in the present study are available upon reasonable request

### Appendix A. Baseline demographics of full Chasing COVID cohort (C3) and comparison to study cohort

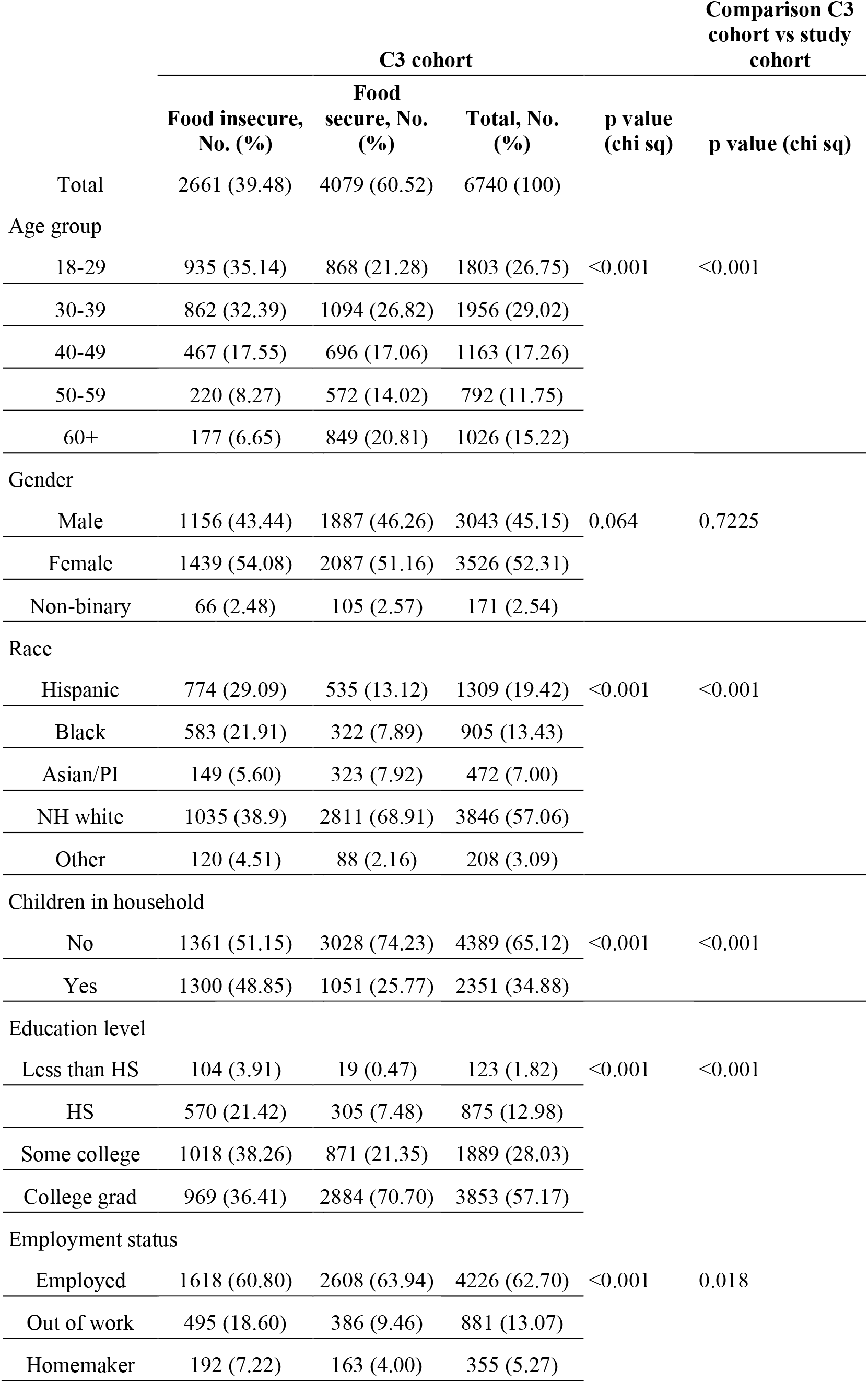
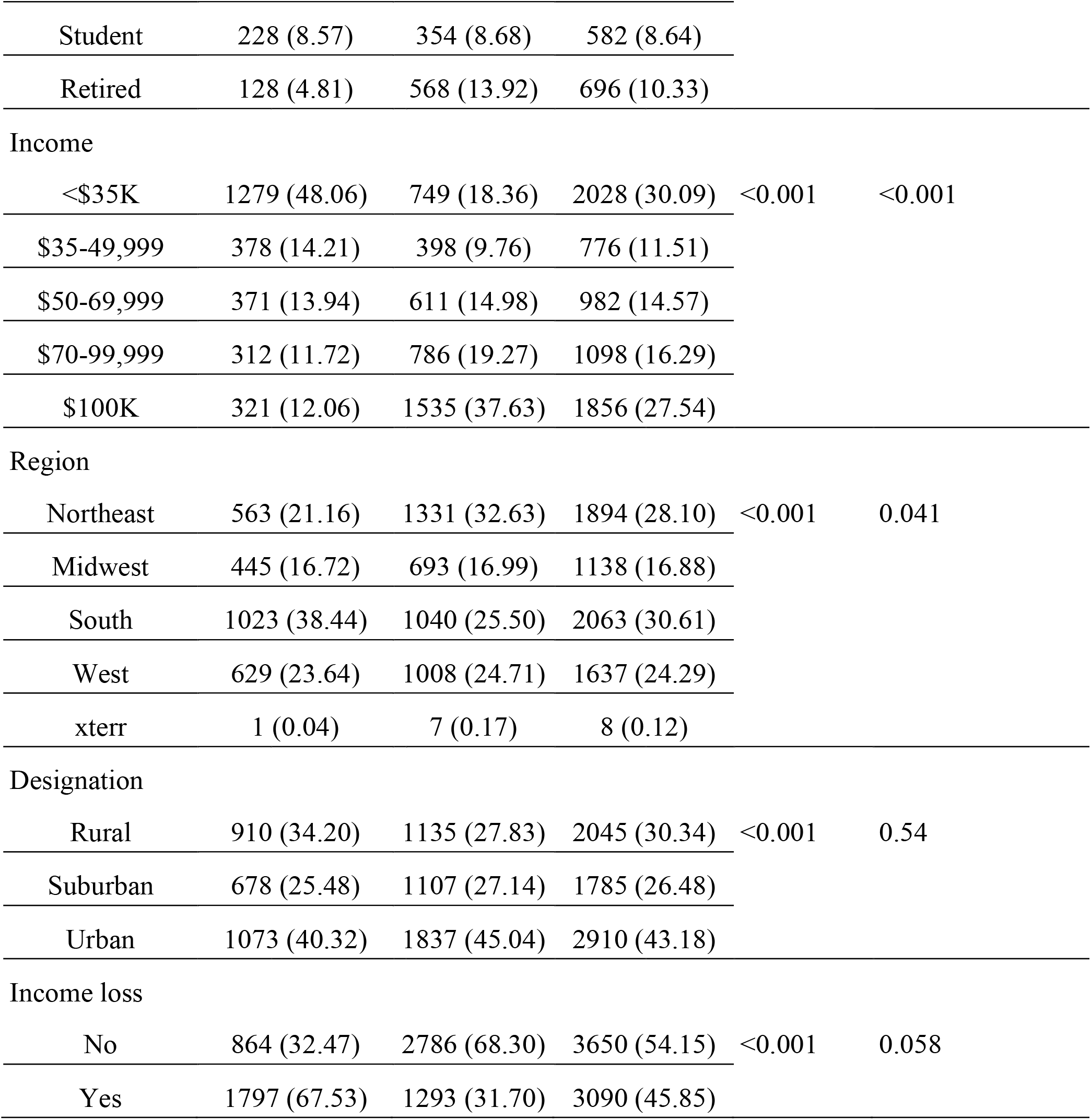

|  | C3 cohort |  |  |  | Comparison C3 cohort vs study cohort |
| --- | --- | --- | --- | --- | --- |
|  | Food insecure, No. (%) | Food secure, No. (%) | Total, No. (%) | p value (chi sq) | p value (chi sq) |
| Total | 2661 (39.48) | 4079 (60.52) | 6740 (100) |  |  |
| Age group |  |  |  |  |  |
| 18-29 | 935 (35.14) | 868 (21.28) | 1803 (26.75) | <0.001 | <0.001 |
| 30-39 | 862 (32.39) | 1094 (26.82) | 1956 (29.02) |  |  |
| 40-49 | 467 (17.55) | 696 (17.06) | 1163 (17.26) |  |  |
| 50-59 | 220 (8.27) | 572 (14.02) | 792 (11.75) |  |  |
| 60+ | 177 (6.65) | 849 (20.81) | 1026 (15.22) |  |  |
| Gender |  |  |  |  |  |
| Male | 1156 (43.44) | 1887 (46.26) | 3043 (45.15) | 0.064 | 0.7225 |
| Female | 1439 (54.08) | 2087 (51.16) | 3526 (52.31) |  |  |
| Non-binary | 66 (2.48) | 105 (2.57) | 171 (2.54) |  |  |
| Race |  |  |  |  |  |
| Hispanic | 774 (29.09) | 535 (13.12) | 1309 (19.42) | <0.001 | <0.001 |
| Black | 583 (21.91) | 322 (7.89) | 905 (13.43) |  |  |
| Asian/PI | 149 (5.60) | 323 (7.92) | 472 (7.00) |  |  |
| NH white | 1035 (38.9) | 2811 (68.91) | 3846 (57.06) |  |  |
| Other | 120 (4.51) | 88 (2.16) | 208 (3.09) |  |  |
| Children in household |  |  |  |  |  |
| No | 1361 (51.15) | 3028 (74.23) | 4389 (65.12) | <0.001 | <0.001 |
| Yes | 1300 (48.85) | 1051 (25.77) | 2351 (34.88) |  |  |
| Education level |  |  |  |  |  |
| Less than HS | 104 (3.91) | 19 (0.47) | 123 (1.82) | <0.001 | <0.001 |
| HS | 570 (21.42) | 305 (7.48) | 875 (12.98) |  |  |
| Some college | 1018 (38.26) | 871 (21.35) | 1889 (28.03) |  |  |
| College grad | 969 (36.41) | 2884 (70.70) | 3853 (57.17) |  |  |
| Employment status |  |  |  |  |  |
| Employed | 1618 (60.80) | 2608 (63.94) | 4226 (62.70) | <0.001 | 0.018 |
| Out of work | 495 (18.60) | 386 (9.46) | 881 (13.07) |  |  |
| Homemaker | 192 (7.22) | 163 (4.00) | 355 (5.27) |  |  |
| Student | 228 (8.57) | 354 (8.68) | 582 (8.64) |  |  |
| Retired | 128 (4.81) | 568 (13.92) | 696 (10.33) |  |  |
| Income |  |  |  |  |  |
| <\$35K | 1279 (48.06) | 749 (18.36) | 2028 (30.09) | <0.001 | <0.001 |
| \$35-49,999 | 378 (14.21) | 398 (9.76) | 776 (11.51) | | |
| \$50-69,999 | 371 (13.94) | 611 (14.98) | 982 (14.57) | | |
| \$70-99,999 | 312 (11.72) | 786 (19.27) | 1098 (16.29) | | |
| \$100K | 321 (12.06) | 1535 (37.63) | 1856 (27.54) | | |
| Region |  |  |  |  |  |
| Northeast | 563 (21.16) | 1331 (32.63) | 1894 (28.10) | <0.001 | 0.041 |
| Midwest | 445 (16.72) | 693 (16.99) | 1138 (16.88) |  |  |
| South | 1023 (38.44) | 1040 (25.50) | 2063 (30.61) |  |  |
| West | 629 (23.64) | 1008 (24.71) | 1637 (24.29) |  |  |
| xterr | 1 (0.04) | 7 (0.17) | 8 (0.12) |  |  |
| Designation |  |  |  |  |  |
| Rural | 910 (34.20) | 1135 (27.83) | 2045 (30.34) | <0.001 | 0.54 |
| Suburban | 678 (25.48) | 1107 (27.14) | 1785 (26.48) |  |  |
| Urban | 1073 (40.32) | 1837 (45.04) | 2910 (43.18) |  |  |
| Income loss |  |  |  |  |  |
| No | 864 (32.47) | 2786 (68.30) | 3650 (54.15) | <0.001 | 0.058 |
| Yes | 1797 (67.53) | 1293 (31.70) | 3090 (45.85) |  |  |

### Appendix B. Correlation of demographic factors with food insecurity among full Chasing COVID cohort (C3)

|  | Univariable Analysis |  | Multivariable Analysis |  |
| --- | --- | --- | --- | --- |
|  | OR (95% CI) | P value | OR (95% CI) | P value |
| Age group |  |  |  |  |
| 18-29 (ref) | - | - | - | - |
| 30-39 | 0.73 (0.64, 0.83) | <0.001 | 0.85 (0.72, 1.00) | <0.001 |
| 40-49 | 0.62 (0.54, 0.72) | <0.001 | 0.74 (0.61, 0.90) | 0.02 |
| 50-59 | 0.36 (0.30, 0.43) | <0.001 | 0.45 (0.36, 0.57) | <0.001 |
| 60+ | 0.19 (0.16, 0.23) | <0.001 | 0.34 (0.26, 0.45) | <0.001 |
| Gender |  |  |  |  |
| Male (ref) | - | - | - | - |
| Female | 1.13 (1.02, 1.25) | 0.023 | - | - |
| Non-binary | 1.03 (0.75, 1.41) | 0.84 | - | - |
| Race |  |  |  |  |
| NH white (ref) | - | - | - | - |
| Hispanic | 3.93 (3.45, 4.49) | <0.001 | 2.07 (1.77, 2.44) | 0.002 |
| Black | 4.92 (4.22, 5.74) | <0.001 | 2.44 (2.02, 2.94) | <0.001 |
| Asian/PI | 1.26 (1.02, 1.54) | <0.001 | 1.16 (0.91, 1.48) | <0.001 |
| Other | 3.71 (2.79, 4.93) | <0.001 | 2.25 (1.60, 3.17) | 0.03 |
| Children in household |  |  |  |  |
| No (ref) | - | - | - | - |
| Yes | 2.76 (2.48, 3.06) | <0.001 | 2.21 (1.94, 2.53) | <0.001 |
| Education level |  |  |  |  |
| Less than HS (ref) | - | - | - | - |
| HS | 0.34 (0.21, 0.57) | <0.001 | 0.27 (0.15, 0.48) | 0.38 |
| Some college | 0.21 (0.13, 0.35) | 0.009 | 0.23 (0.13, 0.41) | 0.004 |
| College grad | 0.06 (0.04, 0.10) | <0.001 | 0.12 (0.07, 0.21) | <0.001 |
| Employment status |  |  |  |  |
| Employed (ref) | - | - | - | - |
| Out of work | 2.07 (1.79, 2.39) | <0.001 | 1.05 (0.87, 1.26) | 0.02 |
| Homemaker | 1.90 (1.53, 2.36) | <0.001 | 1.04 (0.78, 1.38) | 0.12 |
| Student | 1.04 (0.87, 1.24) | 0.60 | 0.47 (0.37, 0.59) | <0.001 |
| Retired | 0.36 (0.29, 0.44) | <0.001 | 0.95 (0.70, 1.29) | 0.47 |
| Income |  |  |  |  |
| <\$35K (ref) | - | - | - | - |
| \$35-49,999 | 0.56 (0.47, 0.66) | <0.001 | 0.66 (0.54, 0.80) | <0.001 |
| \$50-69,999 | 0.36 (0.30, 0.42) | 0.97 | 0.43 (0.36, 0.52) | 0.36 |
| \$70-99,999 | 0.23 (0.20, 0.27) | <0.001 | 0.34 (0.28, 0.41) | <0.001 |
| \$100K | 0.12 (0.11, 0.14) | <0.001 | 0.21 (0.18, 0.26) | <0.001 |
| Region |  |  |  |  |
| Northeast (ref) | - | - | - | - |
| Midwest | 1.52 (1.30, 1.77) | 0.91 | 1.07 (0.89, 1.30) | 0.76 |
| South | 2.33 (2.04, 2.65) | <0.001 | 1.33 (1.13, 1.57) | <0.001 |
| West | 1.47 (1.28, 1.69) | 0.56 | 1.00 (0.84, 1.19) | 0.11 |
| Designation |  |  |  |  |
| Urban | - | - | - | - |
| Rural | 1.37 (1.22, 1.54) | <0.001 | 1.19 (1.03, 1.38) | 0.04 |
| Suburban | 1.05 (0.93, 1.18) | 0.05 | 1.07 (0.92, 1.24) | 0.74 |
| Income loss |  |  |  |  |
| No (ref) | - | - | - | - |
| Yes | 4.49 (4.04, 4.98) | <0.001 | 3.72 (3.29, 4.22) | <0.001 |

